# ToxPi*GIS Toolkit: Creating, viewing, and sharing integrative visualizations for geospatial data using ArcGIS

**DOI:** 10.1101/2021.10.08.21264756

**Authors:** Jonathon Fleming, Skylar W. Marvel, Alison A. Motsinger-Reif, David M. Reif

## Abstract

**Background:** Presenting a comprehensive picture of geographic data comprising multiple factors is an inherently integrative undertaking. Visualizing such data in an interactive form is essential for public sharing and geographic information systems (GIS) analysis. The Toxicological Prioritization Index (ToxPi) framework has been used as an integrative model layered atop geospatial data, and its deployment within the dynamic ArcGIS universe would open up powerful new avenues for sophisticated, interactive GIS analysis.

**Objective:** We propose an actively developed suite of software, the ToxPi*GIS Toolkit, for creating, viewing, sharing, and analyzing interactive ToxPi figures in ArcGIS.

**Methods:** The ToxPi*GIS Toolkit is a collection of methods for creating interactive feature layers that contain ToxPi diagrams. It currently includes an ArcGIS Toolbox (*ToxPiToolbox*.*tbx*) for drawing geographically located ToxPi diagrams onto a feature layer, a collection of modular Python scripts that create predesigned layer files containing ToxPi feature layers from the command line, and a collection of Python routines for useful data manipulation and preprocessing. We present workflows documenting ToxPi feature layer creation, sharing, and embedding for both novice and advanced users looking for additional customizability.

**Results:** Map visualizations created with the ToxPi*GIS Toolkit can be made freely available on public URLs, allowing users without ArcGIS Pro access or expertise to view and interact with them. Novice users with ArcGIS Pro access can create *de novo* custom maps, and advanced users can exploit additional customization options. The ArcGIS Toolbox provides a simple means for generating ToxPi feature layers. We illustrate its usage with current COVID-19 data to compare drivers of pandemic vulnerability in counties across the United States.

**Significance:** Development of new features, which will advance the interests of the scientific community in many fields, is ongoing for the ToxPi*GIS Toolkit, which can be accessed from www.toxpi.org.

**Impact Statement:** Presenting a comprehensive picture of geographic data comprising multiple factors is an inherently integrative undertaking. Visualizing this data in an interactive form is essential for public sharing and geographic analysis. The ToxPi framework provides such integration, and ArcGIS offers interactive geographic mapping capability, but, so far, producing ToxPi figures in ArcGIS maps has not been possible. We propose the ToxPi*ArcGIS Toolkit, which enables the generation of ArcGIS feature layers that include interactive ToxPi figures. Further, we document the living code repository created for this method and outline workflows for sharing, creating, and embedding maps within a web dashboard.

**Availability and Implementation:** All applications, usage instructions, sample data, example visualizations, and open-source code are freely available from a dedicated GitHub page linked from www.toxpi.org. ArcGIS Pro can be obtained at https://www.esri.com/en-us/arcgis/products/arcgis-pro/overview.

## Introduction

Geographic data are used to support decisions and inform analysis at many scales, from neighborhoods and larger communities to states and nations. Combined with sophisticated GIS tools, these data can be a powerful means of communicating issues affecting communities and their drivers and can act as a catalyst for change. Because of the complexity of places and their underlying environments, data from a single source is often insufficient for accurately communicating a geographical area’s issues and the drivers of problems such as health disparities. To remedy this, summary quantitative layers, or scores, can be created by aggregating multiple data sources. While these layers or scores convey the combined effect of numerous factors on a population, there is typically no mechanism to account for the contributions of individual factors to the overall score. For a truly comprehensive picture of geographic data comprising multiple factors, visual analytics are needed that present both integrated scores and the contributions of the distinct factors driving these scores.

One such integrative visual analytic is the Toxicological Prioritization Index (ToxPi) framework, which displays results as radar charts composed of “slices” (ref. 1). The flexible ToxPi framework accepts continuous and ordinal data for integration as sets of related, user-defined factors into slice-wise and overall scores (see Figure 1). The framework is integrated into the ToxPi GUI, a Java application with an easy-to-use graphical interface for analyzing and visualizing diverse data into ToxPi profiles (www.toxpi.org) (ref. 2).

**Figure 1.**
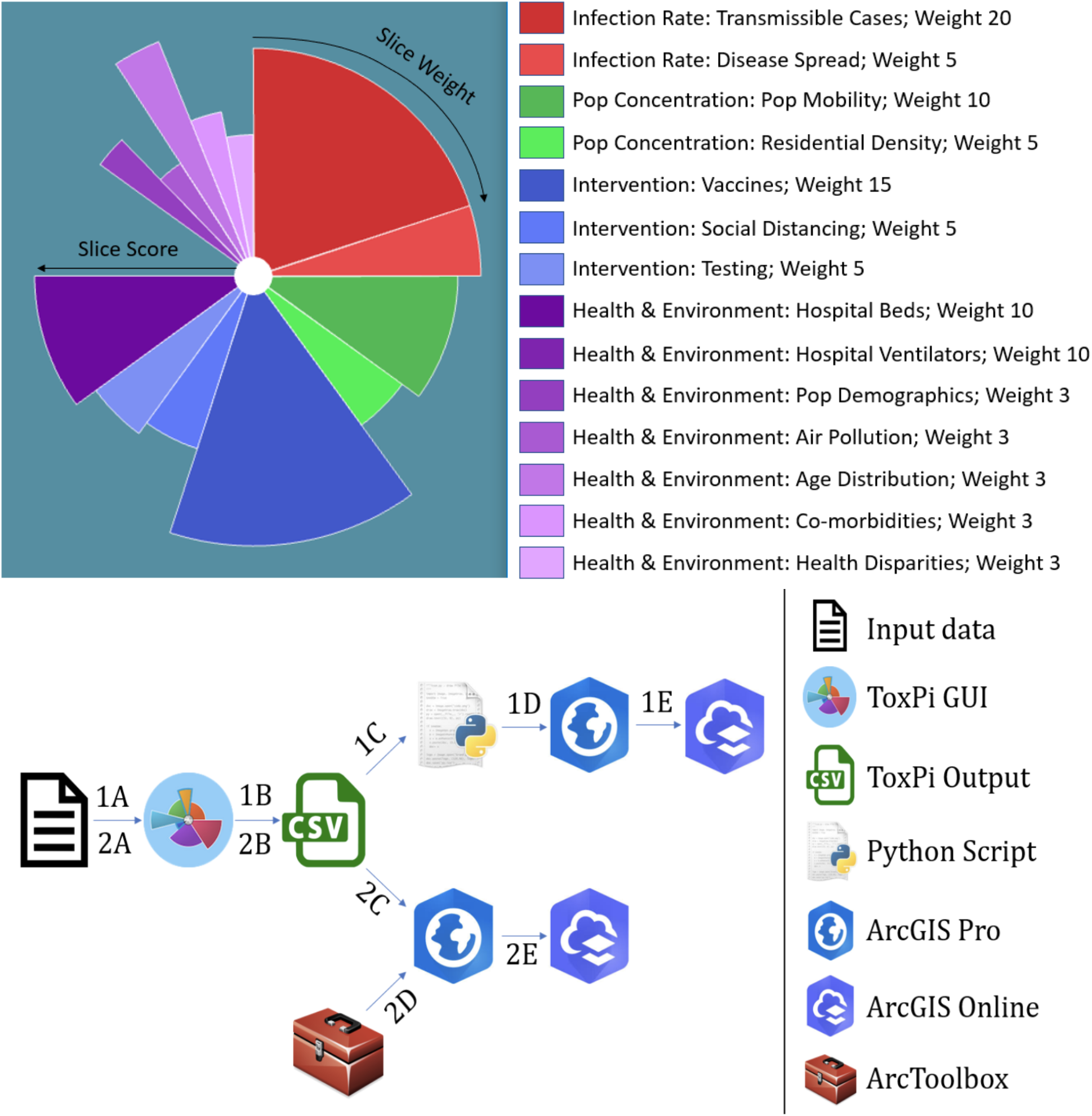
(Top) Overview of the ToxPi framework for COVID-19 vulnerability in the new ‘PVI with Vaccine Model’ with four domains (color families) and 14 subdomains (slices) and vaccine data. (Bottom) Schematic of the ToxPi*GIS Toolkit workflow Methods 1 and 2, from raw data to map sharing.

The ToxPi framework provides an integrative model layer atop geospatial data, for a compact summary of the factors driving differences in analysis results among regions. It has been deployed within custom-built dashboards created to assess vulnerability to COVID-19 at the county level across the United States (ref. 3), to compare hurricane susceptibility at the census-tract level in the greater Houston area (ref. 4), and to visualize the interplay of stressors related to children’s environmental health across North Carolina (ref. 5). The ToxPi*GIS web application can also be used in tandem with ArcGIS tools/dashboards for parallel analysis (ref. 6). These applications demonstrate the utility of graphics that integrate geographic and multivariate data for addressing the disparate effects of various drivers on vulnerability and susceptibility in different locations.

ArcGIS Pro provides advanced capabilities for geospatial data analysis and map production, and ArcGIS Online is a user-friendly interface that can be used to share these maps. Despite these advantages, so far, there has been no tool for integrating interactive ToxPi figures within an ArcGIS map. To solve this issue, we developed an ArcGIS Pro Toolbox that draws location-based ToxPi figures as feature layers for placement on an ArcGIS map. The use of ArcGIS tools often requires preliminary data manipulation and several sequential, often complicated geoprocessing steps for feature layer preparation, making map creation a daunting task for inexperienced ArcGIS users. To simplify map creation, we developed a custom Python script, *ToxPi_creation*.*py*, that handles all data preparation, generates a layer file containing the drawn ToxPi figures, and displays the layer based on the user’s choices in the ToxPi GUI. We combined the toolbox and the Python script to create the ToxPi*GIS Toolkit, a continuously developed repository of software for integrating ToxPi statistics and the results of geospatial analysis.

The applications are detailed for those without ArcGIS Pro access, novice users with ArcGIS Pro access but limited expertise, and advanced users looking to create customized, integrated ArcGIS workflows and distributable dashboards. Novice users of ArcGIS Pro can generate shareable maps with limited knowledge of the software, and advanced users can use the Toolkit to generate ToxPi feature layers with options for customization, integration, and distribution. Created maps can be made freely available on public URLs so that ArcGIS Pro access is not required to view and interact with them. Here, we illustrate applications of the ToxPi*GIS Toolkit using publicly available COVID-19 data to compare the pandemic vulnerability of counties across the United States. All applications, usage instructions, sample data, example visualizations, and open-source code are freely available from a dedicated GitHub page linked on www.toxpi.org.

## Methods

The ToxPi*GIS Toolkit is an addendum of methods to be used alongside ToxPi GUI, a free, platform-independent Java application for recombining diverse source data into ToxPi profiles (www.toxpi.org). The toolbox methods proposed in this paper can be used to produce interactive feature layers containing ToxPi diagrams for map creation and visualization.

### Toxicological Priority Index (ToxPi)

The ToxPi framework provides a method for transparently integrating and visualizing data across disparate information domains and is often used to determine risk values for the data being analyzed. Data that are not normally compared are combined into a data matrix comprising various data domains, or slice categories, with varying weights that represent the different data categories. Each slice category, represented by a color scheme, can then be separated into subdomains. The data matrix is then analyzed to produce a score ranging from 0 to 1 for each slice that represents a data record’s (e.g. a single county) score (e.g. vulnerability to COVID-19 driven by this subset of factors). The slices are then combined into a normalized overall score that represents the record’s total risk from all slices (i.e. the entire ToxPi profile for a county). These scores are displayed in a ToxPi diagram (shown at the top of *Figure 1*) for each record. Although these visualizations allow the easy comparison of records, when integrated with geospatial data, a large number of figures, which cannot be quickly compared, is generated. Further, these figures do not geographically represent the source of the data or indicate what underlying data was used to generate each slice.

### ToxPi*GIS Toolkit

The ToxPi*GIS Toolkit uses custom Python scripts combined with ArcGIS Pro methods and a custom ArcGIS Pro Toolbox to generate feature layers that contain ToxPi figures drawn at the location where each record was obtained, enabling the geospatial comparison of records. These feature layers can be loaded onto a new map or one with existing data, enabling new comparisons. Furthermore, each slice is drawn as a separate polygon, which allows users to select an individual slice to obtain more information about the feature it represents. These details are displayed in popups that can be easily reconfigured to display important, underlying data used in the analysis. The ToxPi*GIS Toolkit currently includes two suggested methods for generating these feature layers, which are described below. The pipelines for the use cases are outlined at the bottom of *Figure 1*.

### Method 1- Creation of ToxPi feature layers using Python scripts

Method 1 consists of a Python script, *ToxPi_creation*.*py*, that can be run from the Windows command prompt. The output of ToxPi GUI is used as input to produce ToxPi feature layers. The script automates all geoprocessing and data manipulation steps required for ToxPi feature layer production and has only two required parameters—the location of the input data file and the location for the script output. Furthermore, it provides an option to scale the size of ToxPi figures so users can make adjustments based on their data’s scope and density.

The simplicity of this method allows users of all experience levels to quickly and easily produce feature layers that are ready to be shared as a map. These feature layers are saved to a layer file (.lyrx) that can be opened with ArcGIS Pro and shared to ArcGIS Online for public viewing. Users can create a shareable map by opening an output layer file in ArcGIS Pro and selecting the option to publicly share it to ArcGIS Online, allowing anyone with the URL to view and interact with the map. A map owner can also allow users to copy and further analyze the map and its underlying data to produce new, interactive visualizations.

This method is suggested for users who are unfamiliar with ArcGIS Pro as well as those who do not require significant customization and want to bypass manual data and feature class preparation. The script’s input-data formatting requirements are outlined in the Toolkit documentation, and an evolving Utilities folder contains information that can help with these requirements.

### Method 2- Creation of ToxPi feature layers using an ArcGIS Toolbox

Method 2 consists of an ArcGIS Pro Toolbox, *ToxPiToolbox*.*tbx*. This method outputs a feature layer that can be integrated into other analysis pipelines and shared to ArcGIS Online for public viewing. While the method allows more customization of feature layers than the Python script, including the ability to draw a subset of slices and use different coordinate systems, it requires more data preparation. The Toolkit documentation contains an example walkthrough of the steps for data preparation and the creation of ToxPi feature layers. While experienced ArcGIS Pro users can easily customize some layers during preparation, including associating more underlying data with each ToxPi figure, several steps are compulsory for preparing toolbox input data.

Notably, feature classes or layers prepared for the tool must be converted into a projected coordinate system prior to input. *Figure 2* describes the interface for the toolbox and describes each parameter. *Figure 3* provides an example map of local vulnerability to COVID-19 produced by running the python script or toolbox with COVID-19 data.

**Figure 2.**
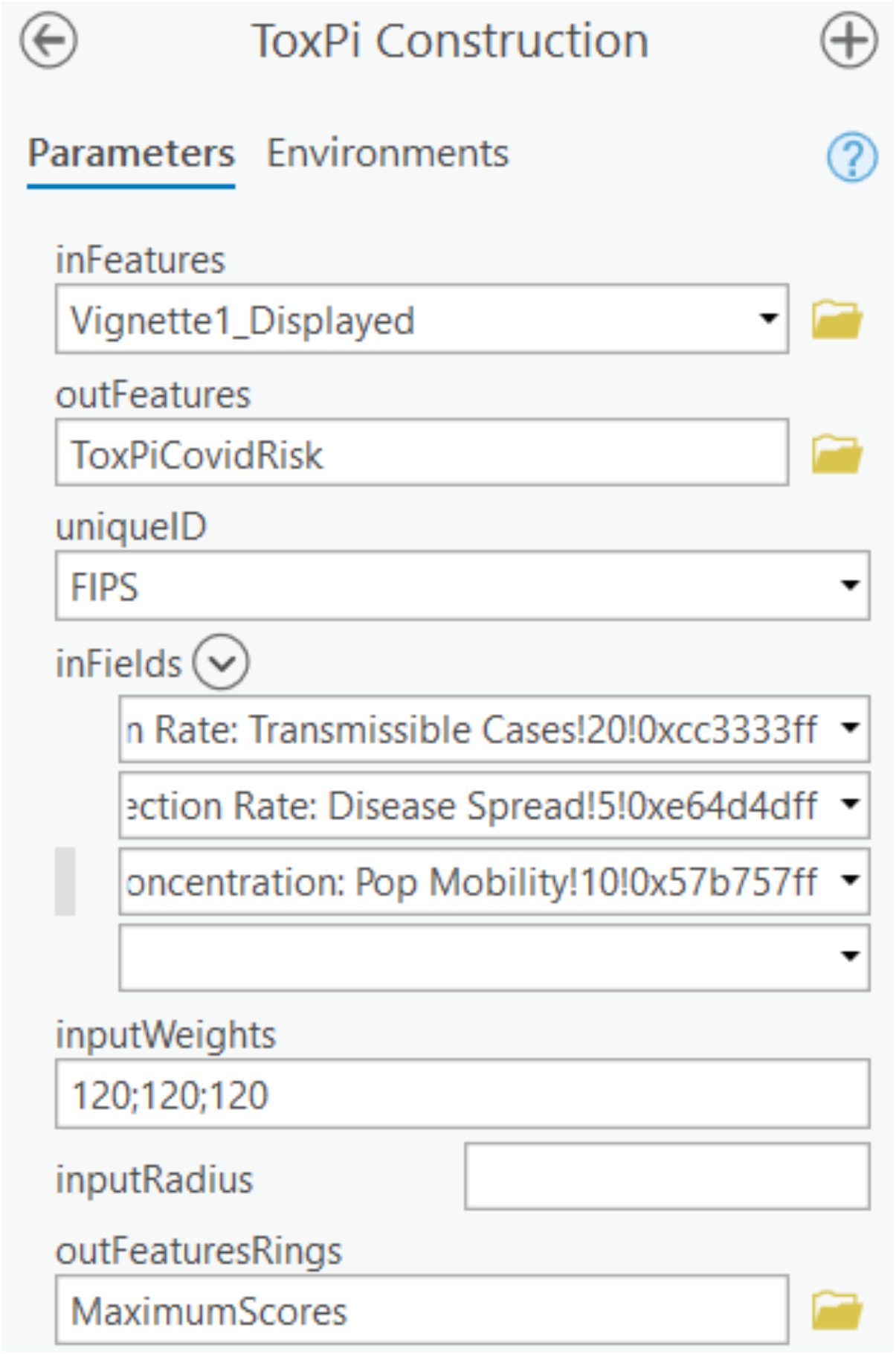
Interface for ToxPi construction using the ToxPi Toolbox. Parameters: inFeatures–the name of the input feature layer; outFeatures–the name of the output feature class; uniqueID–the unique identifier for each data point; inFields–a list of the slices to be included in the ToxPi figures; inputWeights–a string of weights that correspond to the list of fields for determining radial width; inputRadius–optional value to scale ToxPi size; outFeaturesRings–optional name of the maximum radius rings feature layer

**Figure 3.**
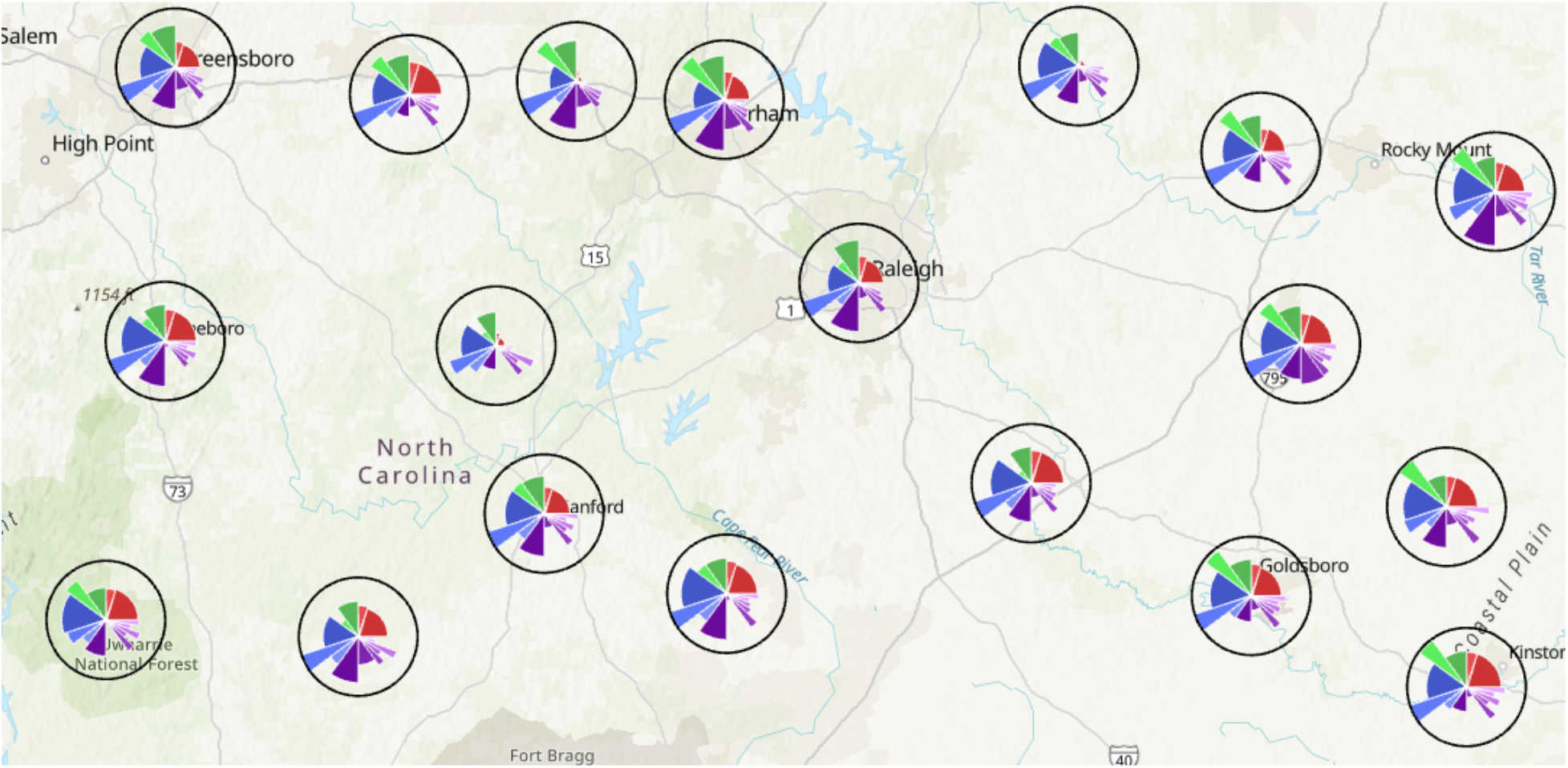
Output of *ToxPi_creation*.*py* for COVID-19 vulnerability data zoomed in on a section of North Carolina

### Advancement and versatility of ToxPi_creation.py

To demonstrate the versatility of the ToxPi*GIS Toolkit and the customization capacity provided by ArcGIS, we added advanced geoprocessing steps and analysis for use with county- or census-tract-level data to the *ToxPi_creation*.*py* script for Method 1. The resulting script, *ToxPi_creation_customized*.*py*, can be run with the same workflow as Method 1 but produces more advanced maps for a specific geographic data type. This method generates extra layers for visualization, including two separately-sized layers of ToxPi features to account for areas of different geographic boundaries, from local to state to regional. These include a local choropleth layer representing the ToxPi score, a state-level ToxPi layer containing the state medians as ToxPi features, and a state-level choropleth layer representing the ToxPi score. *Figure 4* contains static images of a map at different zoom extents that was produced by running the altered script with COVID-19 vulnerability data, thus showing the multilayer capability of the altered script and demonstrating the integration of ToxPi_creation.py with other advanced geoprocessing steps.

**Figure 4.**
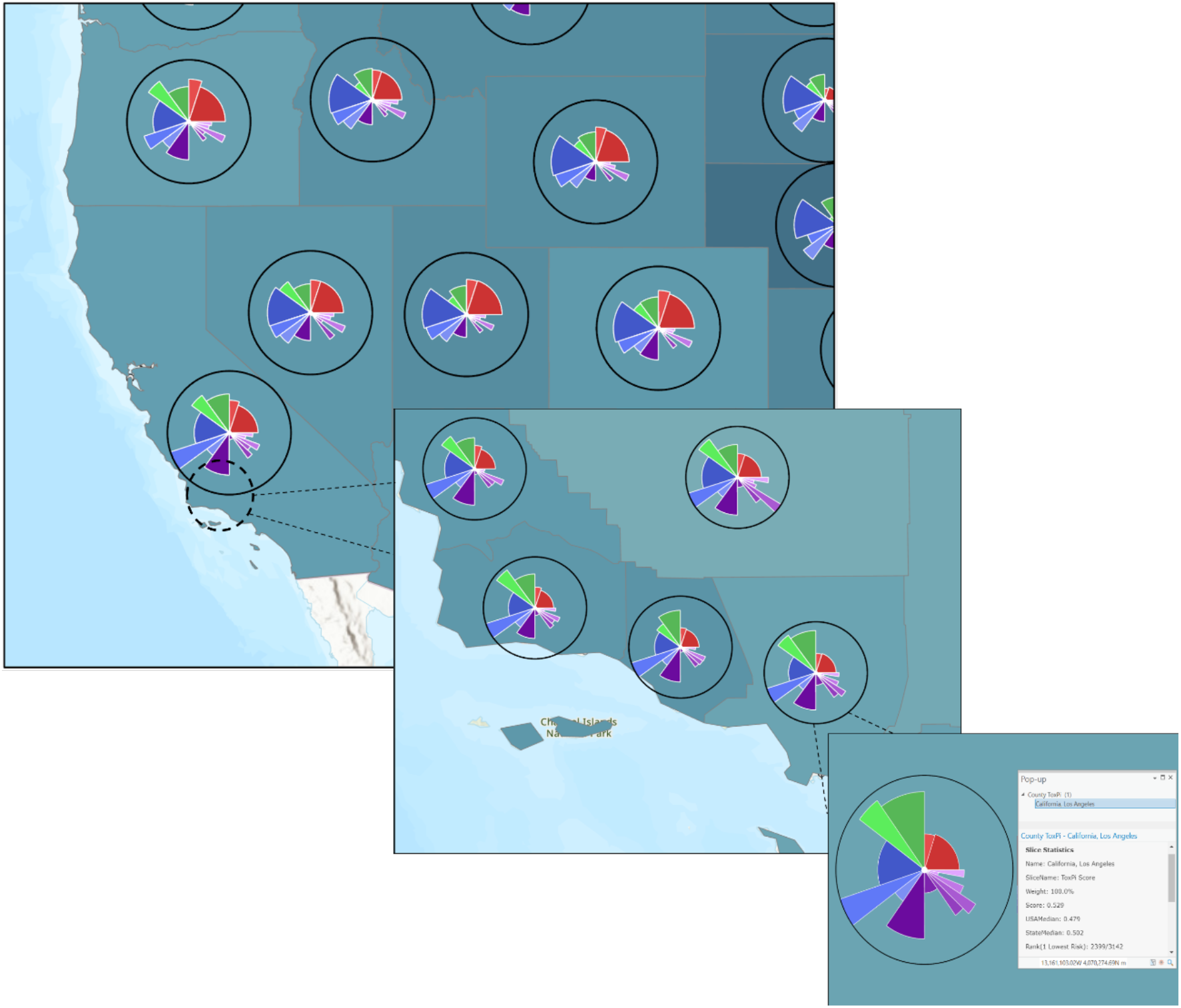
Output of *ToxPi_creation_customized*.*py* with COVID-19 vulnerability data showing the switching of feature layers based on zoom extent. The top left (zoomed out) is a state median feature layer, the middle (zoomed in) is a local ToxPi feature layer, and the bottom right (zoomed in further) is a local ToxPi feature layer that accounts for overlap in high-density areas. A popup with important information appears when selecting a ToxPi feature.

### Data collection

We obtained data for the COVID-19 vulnerability map from the Pandemic Vulnerability Index (PVI) Dashboard at https://covid19pvi.niehs.nih.gov/. While the PVI Dashboard was introduced prior to the availability of vaccines, it now contains a new model that includes slices representing vaccination data available starting January 2021, so we focused on data since that time. In this model, risk components are split into four domains—infection rate, population concentration, intervention measures, and health and environment—that comprise 14 categories. *Figure 1* shows each category’s corresponding weight. This model accounts for vaccine data in the intervention measures domain, allowing for a more accurate representation of COVID-19 risk than previous models. More information about the model, including the underlying components for each slice category, can be found at the National Institutes of Health (NIH) Pandemic Vulnerability Index (PVI) Dashboard details page (https://www.niehs.nih.gov/research/programs/coronavirus/covid19pvi/details/). We used these data to demonstrate the extended capabilities of the ToxPi*GIS Toolkit, including its integration with powerful analysis methods available in ArcGIS and the visualization capabilities of ArcGIS Online.

### New advanced analysis options provided by ArcGIS Pro

To demonstrate the powerful statistical analysis methods ArcGIS Pro provides, we performed hotspot analysis using ToxPi scores and county-level results from the COVID-19 vulnerability model created with *ToxPi_creation_customized*.*py*. We conducted the hotspot analysis in ArcGIS Pro using the Optimized Hotspot Analysis tool with a distance band of 50 miles. Given incident points or weighted features (points or polygons), this tool creates a map of statistically significant hotspots and coldspots using the Getis-Ord Gi* statistic and evaluates the characteristics of the input feature class to produce optimal results. The tool creates a confidence-level bin with a corresponding z-score and p-value for each feature within the input feature class (ref. 7). In this case, each county was placed into a Gi_Bin based on the confidence level that it was clustered in a hotspot or coldspot according to the ToxPi score. By default, seven bins were generated, +/-3, +/-2, +/-1, and 0, where + denotes a hotspot, - demotes a cold spot, 3 denotes a 99% confidence interval, 2 denotes a 95% confidence interval, 1 denotes a 90% confidence interval, and 0 denotes no statistical significance.

The result is a choropleth layer of counties that can easily be used to determine clusters of high and low risk for COVID-19, as shown in *Figure 5*. Zooming in on this layer displays the ToxPi figures, which can be used as a visual reference for further analysis of the factors causing these clusters. Although the analysis was conducted using the overall ToxPi score, it could easily be repeated using a specific slice to allow for more specific statistical analysis of how individual factors compare across a geographic region. To demonstrate some of the powerful hosting and visualization capabilities ArcGIS provides, we joined the hotspot analysis results with raw data and displayed the result in an ArcGIS dashboard for public viewing.

**Figure 5.**
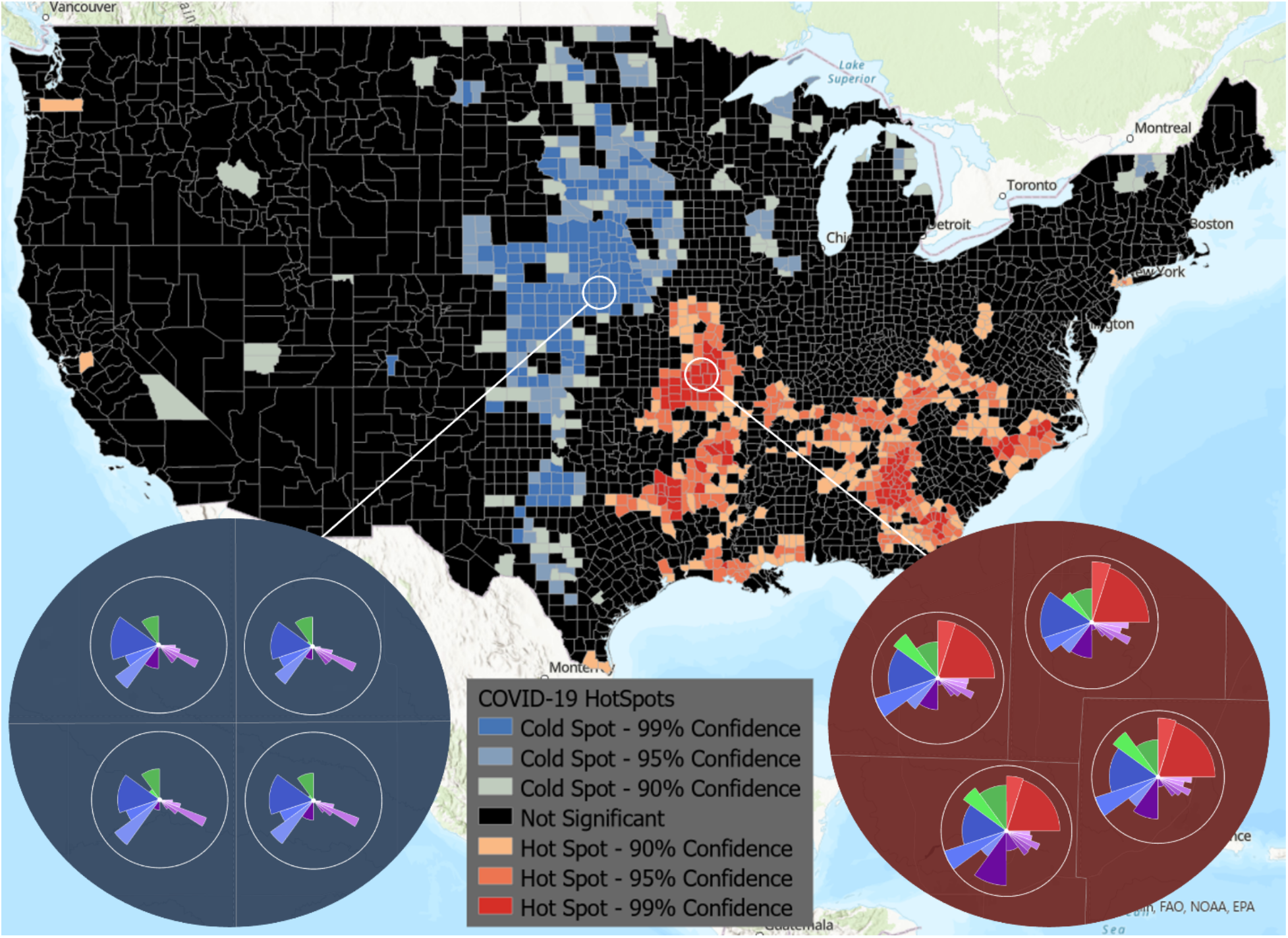
Hotspot analysis provided by ArcGIS integrated with the ToxPi*GIS Toolkit. A national view of hotspots and coldspots is shown, where color represents high-risk (hotspot) and low-risk (coldspot) areas. The two insets show ToxPi images for example coldspot (blue) and hotspot (red) counties with a 99% confidence level, providing an easy means of visually comparing the areas based on slice scores.

## Results

All interactive maps and visualizations are linked from www.toxpi.org to the dedicated Github page. *Figure 3* is a static image, zoomed in on a portion of North Carolina, of the model created with COVID-19 data using *ToxPi_creation*.*py*. This method results in an interactive feature layer of ToxPi figures that visually matches the results of *ToxPiToolbox*.*tbx*. However, minor visualization differences are described in the Toolkit documentation.

*Figure 4* is a static image of the result of the model created by *ToxPi_creation_customized*.*py* with COVID-19 data. The figure shows local to regional to state displays that aggregate data by user-selected zoom extent. Because of the interactive nature of these diagrams, users can select individual slices to learn more about the risk factors they represent, enabling quick analysis of the factors driving COVID-19 risk at different geographic scales of interest.

*Figure 5* is a static image of hotspot analysis results and shows that cold spots are primarily clustered in Nebraska and the edges of South Dakota and Kansas. Hotspots are primarily clustered in Missouri, the border of Texas, Arkansas, Georgia, and Tennessee. The zoomed-in views show the ToxPi feature layer for an example hotspot and a coldspot, allowing direct visual comparison of the different factors affecting the regions. The ToxPi radar charts clearly show that transmissible cases, disease spread, residential density, social distancing, population demographics, and health disparities are highly different between the hotspot and coldspot.

*Figure 6* is a static image of hotspot analysis results displayed on a publicly viewable dashboard. Important statistics can be easily retrieved for a county of interest. The flexible customization of this powerful visualization technique enables users to tell a story about a location based on a dataset. The figure shows COVID-19 statistics, including cases, deaths, sickness scores, and disease spread rate for Greene County, Missouri, a major hotspot.

**Figure 6.**
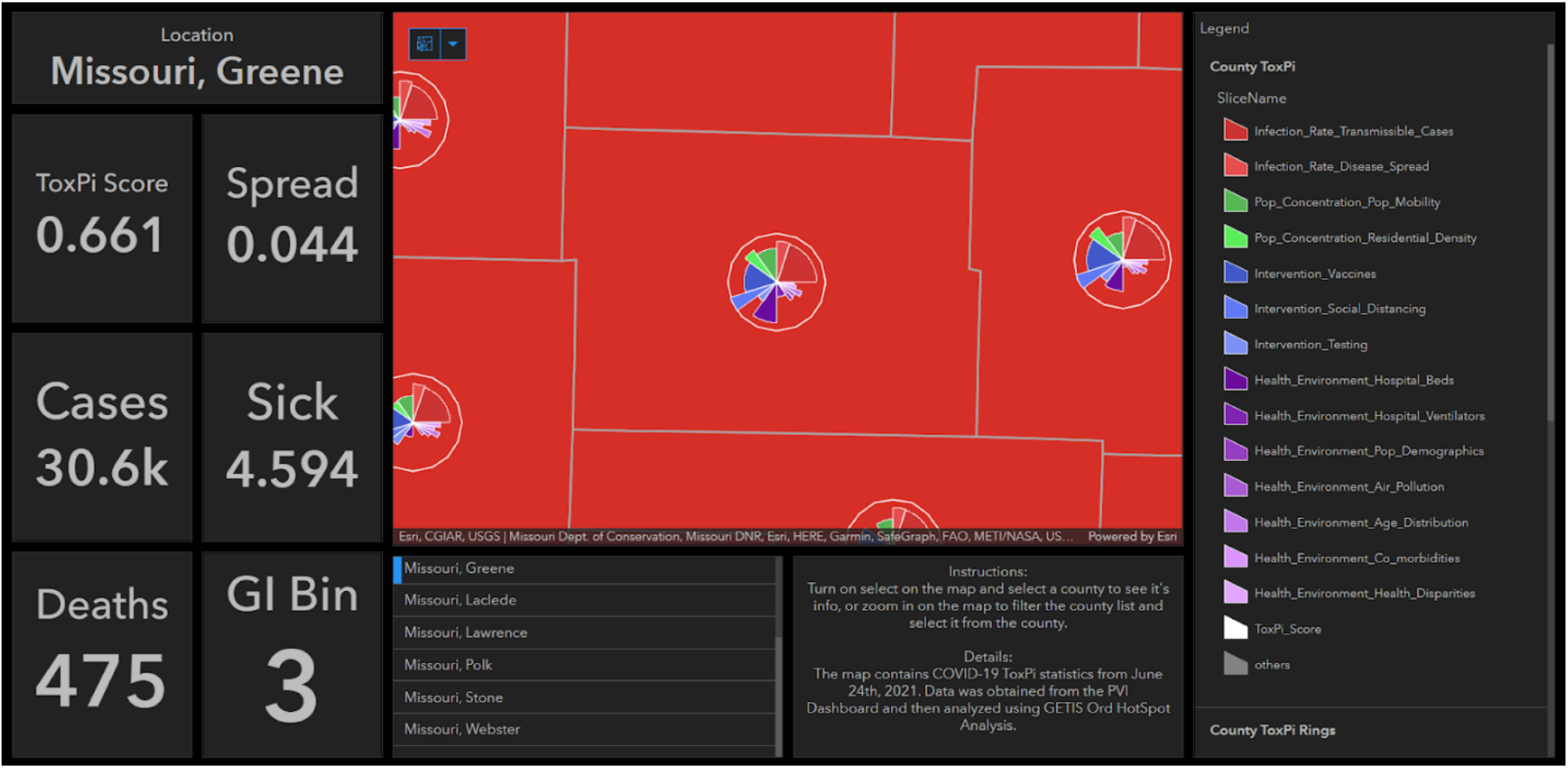
Example dashboard created by joining the hotspot analysis results with the raw data in an ArcGIS Online Dashboard and configuring panels to show important statistics for each county. By selecting a county, users can see its ToxPi score, disease spread rate, sickness score, number of cases and deaths, and the Gi_Bin it was placed in to determine the confidence level of its status as a hotspot or coldspot.

## Discussion

We developed two new methods using ArcGIS Pro to create feature layers containing ToxPi diagrams. The first method comprises a Python script, *ToxPi_creation*.*py*, that is user-friendly and suitable for ArcGIS users of all experience levels as well as those who want to produce a feature layer containing only ToxPi figures. The script automates the required geoprocessing steps, so after loading the ArcGIS Pro environment, users simply run the script with the results of the ToxPi GUI as input. Further, as the script uses the underlying code for the toolbox, users are not required to download and add the toolbox to the proper environment, making it especially accessible for inexperienced ArcGIS users. Because the script is open source, experienced Python users can easily access both the toolbox code and the code used for data preparation and thus develop new related methods and analysis procedures. This flexibility opens the door for new advancements and the creation of complex workflows that will benefit many fields.

The second method comprises an ArcGIS Pro Toolbox, *ToxPiToolbox*.*tbx*, and is recommended for advanced ArcGIS users due to the geoprocessing steps required for data preparation. The method allows customization of the output feature layer and can be easily integrated into existing workflows for ArcGIS models. It is extremely useful as a means for the further advancement of analysis and visualization in existing workflows, as there is no current method for generating ToxPi feature layers in ArcGIS maps.

The *ToxPi_creation_customized*.*py* script is an example of potential user alterations to *ToxPi_creation*.*py*. The script uses both geoprocessing procedures and Python analysis to create advanced maps and is useful for county- and census-tract-level data. Methods such as using joins and altering the zoom extent for feature layers are useful for creating interactive maps with multiple feature layers that show varying levels of data. The choice of COVID-19 data as an example use case of this script is a relatable demonstration and a useful visualization of data of interest to the general public.

Using COVID-19 data, results from the powerful hotspot analysis option in ArcGIS revealed that significant high-risk clusters are located in Missouri, the border of Texas, Arkansas, Georgia, and Tennessee. Combined with the ToxPi*GIS Toolkit, this method enables the easy visualization of factors affecting high-risk areas. The results can help with decisions regarding how resources should be allocated in counties to address the health disparities contributing to their vulnerability to COVID-19. Although the results displayed here are for a selected time period (June 2021), the intention was to demonstrate a topical use case rich in public data for how the ToxPi*GIS Toolkit can be coupled with ArcGIS analysis methods.

ArcGIS dashboards represent powerful ArcGIS customizations that enable visualization to portray the story behind data. A dashboard offers many advantages over a stand-alone map, namely customization and the ability to include extra analysis to indicate the importance of certain information in a particular context. Visualization options that include web applications and story maps allow for further versatility based on a user’s needs.

The ToxPi*GIS Toolkit, as well as instructions for its use, are freely available on a dedicated GitHub page. Development of this living software Toolkit will continue, with the objective of developing new methods that integrate ToxPi statistics. We provide open-source code to enable users to create new methods, with the objective of advancing the interests of the scientific community. All example visualizations and the dedicated GitHub page are linked at www.toxpi.org, which will be regularly updated with advancements related to ToxPi statistics. Future goals are to develop options for scaling and filtering ToxPi figures, methods for separating overlapping ToxPi figures, the ability to integrate timescale data, and the direct provision of map creation and analysis options on ArcGIS Online website for users without ArcGIS Pro access.

## Data Availability

All applications, usage instructions, sample data, example visualizations, and opensource code are freely available from a dedicated GitHub page linked from www.toxpi.org.

## Acknowledgments

We thank the PVI project team for data provision and the Baker Lab at NCSU for software testing. We also thank Dan Schmitt and Logan Wenzel for their expertise in implementation. This research was supported by the National Institutes of Health (NIH) grant awards ES030007 and ES025128, and by intramural funds from the National Institute of Environmental Health Sciences and the National Institute for Allergy and Infectious Diseases.

## SUPPLEMENTAL and OTHER OPTIONAL MATERIAL

All applications, usage instructions, sample data, example visualizations, and open-source code are freely available from a dedicated GitHub page linked from www.toxpi.org. Direct links to specific elements described above are provided here.

ToxPi*GIS Toolkit Github

https://github.com/Jonathon-Fleming/ToxPi-GIS

Vignette1: *ToxPi_creation*.*py* Demonstration

https://ncsu.maps.arcgis.com/home/item.html?id=7c0365b3f75949369b46c07ae4ecf10c

Vignette2: *ToxPi_creation_customized*.*py* Demonstration

https://ncsu.maps.arcgis.com/home/item.html?id=1518637a0b454036a3d0d2fc8239ff08

HotSpot/Dashboard Demonstration

https://ncsu.maps.arcgis.com/home/item.html?id=022416cbc74d430691ad7d2a4cbec229

